# Evaluation of the Clinical Impact of Different Telemedicine Practices in Intensive Care Units: a Stepped-wedge Cluster Randomized Clinical Trial (TELESCOPE 2): study protocol

**DOI:** 10.64898/2026.01.13.26344051

**Authors:** Adriano José Pereira, Bruna Gomes Barbeiro, Tiago Mendonça dos Santos, Maura Cristina dos Santos, Thiago Domingos Corrêa, Alexandre Biasi Cavalcanti, Ary Serpa Neto, Carlos Henrique Sartorato Pedrotti, Fernando Zampieri, Guilherme de Paula Pinto Schettino, Jorge Ibrain Figueira Salluh, Leandro Utino Taniguchi, Leonardo José Rolim Ferraz, Luciano Cesar Azevedo, Otávio Berwanger, Regis Goulart Rosa, Renata Albaladejo Morbeck, Rodrigo Biondi, Suzana Margareth Lobo, Renato Carneiro de Freitas Chaves, Otavio Ranzani

**Affiliations:** Einstein Hospital Israelita, São Paulo, SP, Brazil; Brazilian Research in Intensive Care Network (BRICNet), São Paulo, Brazil; Insper Institute of Education and Research, São Paulo, SP, Brazil; Instituto de Pesquisa do Hcor, São Paulo, SP, Brazil; Australian and New Zealand Intensive Care Research Centre (ANZIC-RC), Monash University, Melbourne, Australia; Universidade Federal do Rio de Janeiro, Rio de Janeiro, RJ, Brazil; Hospital das Clínicas FMUSP, Universidade de São Paulo, São Paulo, Brazil; Hospital Sírio Libanês, São Paulo, SP, Brazil; Hospital Moinhos de Vento, Porto Alegre, RS, Brazil; Hospital Brasília - Unidade Lago Sul (Rede Américas), Brasília, DF, Brazil; Hospital de Base de São José do Rio Preto, São José do Rio Preto, SP, Brazil; Hospital Santa Luzia, Luziânia, GO, Brazil; Massachusetts Institute of Technology, Cambridge, MA, United States; DataHealth Lab, Institut de Recerca Sant Pau (IR SANT PAU), Barcelona, Spain

**Keywords:** telemedicine, critical care, intensive care units, quality improvement, clinical outcomes

## Abstract

**BACKGROUND:** The safety and feasibility of telemedicine in intensive care units (ICU) are well established. However, whether tele-ICU exerts a measurable impact on clinically relevant outcomes remains uncertain.

**METHODS AND ANALYSIS:** The TELESCOPE 2 is a multicenter, open-label, stepped-wedge cluster randomized controlled trial including 25 ICUs in Brazil from January 2024 to January 2026. In a stepped-wedge assignment, ICUs will be randomized and allocated into one of five sequences. All ICUs will receive the interventions staggered at different times. All adult patients admitted in participant ICUs will be eligible for inclusion in the study. Admissions for non-medical reasons, and patients previously included in the TELESCOPE 2 study will be excluded. The trial intervention is multifaceted, comprising three components delivered in combination, via telemedicine: i) daily multidisciplinary rounds led by board-certified intensive care physicians; ii) coordinated care by a multidisciplinary team, including nurses, physiotherapists, and clinical pharmacists); iii) a management strategy focused on quality improvement and patient safety. The primary outcome is ICU length of stay. Secondary outcomes include ICU mortality, in-hospital mortality, ventilator-free days during the first 28 days, ICU readmission within 48 hours, early reintubation, ventilator-associated events, and accidental extubation rate.

**ETHICS AND DISSEMINATION:** This study was approved by the Institutional Review Board of the Einstein Hospital Israelita (CAAE: 69575123.0.1001.0071), and each participating center’s ethics committee. The study results will be published in peer-reviewed journals and disseminated at international conferences.

**TRIAL REGISTRATION:** Clinicaltrials NCT05960994, Brazilian Registry of Clinical Trials RBR-342wxn9, and Universal Trial Number U1111-1298-9799.

## INTRODUCTION

Telemedicine in intensive care units (Tele-ICU) refers to a wide range of technology-enabled care delivered from remote locations. ^(1–6)^ Tele-ICU integrates secure audio-visual connections and electronic medical records to facilitate remote collaboration between specialized intensive care professionals and on-site ICU staff. ^(1–6)^ Additionally, tele-ICU has the appeal of offering a potential solution to the observed intensivist shortage and to the uneven distribution of specialized intensive care professionals in several countries across the globe. ^(1–6)^

The adoption of tele-ICU has expanded rapidly and is increasingly promoted as a strategy to enhance the quality and consistency of care. ^(6–8)^ Tele-ICU enables remote daily multidisciplinary rounds (DMR) led by an intensive care specialist, provides tools for documenting these rounds, offers second opinions on clinical cases, facilitates consultations with other medical specialties, monitors vital signs in real-time, assists in patient management, and contributes to administrative workflows.^(9)^ Previous non-randomized studies have demonstrated that tele-ICU implementation is associated with reductions in ICU mortality, ^(2, 6–8)^ hospital length of stay, ^(6–8)^ clinical complications, ^(7)^ and faster response to alarms.^(8)^

Despite encouraging evidence from observational studies and systematic review with metanalysis, ^(1–8)^ randomized clinical trials evaluating the impact of tele intensive care systems on patient centered outcomes remain limited. ^(9)^ The TELESCOPE trial, a parallel cluster randomized study conducted in 30 general units in Brazil and enrolling more than 17,000 patients, examined whether remote DMR combined with monthly audit and feedback sessions could reduce intensive care length of stay. ^(9)^ Although the intervention was feasible and safe, it did not reduce the ICU length of stay among critically ill adult patients.

Whether alternative tele-ICU models involving multidisciplinary teams, and management strategies carried out by specialized professionals, could improve outcomes remains unclear. Telescope 2 trial study aims to evaluate whether a multifaceted telemedicine intervention, comprising DMR led by board-certified intensive care physicians, coordinated care from a multidisciplinary team, and a management strategy focused on quality improvement and patient safety, can reduce the ICU length of stay for patients in Brazil.

## METHODS

### Study design and setting

Telescope 2 study is an open-label, national, multicenter, stepped-wedge cluster randomized controlled trial. This study protocol was designed according to the guidelines for Good Clinical Practice and the Declaration of Helsinki and is reported according to the SPIRIT statement. ^(10)^ The study is registered at Clinicaltrials (www.clinicaltrials.gov; trial identification number NCT05960994), the Brazilian Registry of Clinical Trials (https://ensaiosclinicos.gov.br; trial identification number RBR-342wxn9), and the Universal Trial Number is U1111-1298-9799. The main characteristics of the Telescope 2 study are summarized in the Synopsis table (Table 1).

**Table 1.** Synopsis (ClinicalTrials.gov registration, as originally submitted)

### Sites and recruitment

Twenty-five ICUs in Brazil, part of the Brazilian public health system, were recruited for this study. The ICUs were selected from a list of all Brazilian ICUs, provided by the National Council of Municipal Health Secretaries, with support from the of the Brazilian Ministry of Health, to reflect the actual distribution of ICU beds across Brazil’s geographic regions.

### Patient and public involvement

Patients or the public were not involved in the study design.

### Eligibility criteria and inclusion/exclusion criteria

ICUs selected for the study were invited via electronic communication to participate in an interview aimed at assessing their eligibility, using an electronic feasibility assessment questionnaire. All patients admitted to the included ICUs will be eligible for inclusion in the study. The inclusion and exclusion criteria for both ICUs and eligible patients are detailed below.

#### Inclusion criteria for ICUs

- ICU from public or philanthropic hospitals.
- ICUs with a minimum of 7 and a maximum of 20 beds.
- ICU with on-site registered doctors and nurses available 24 hours a day and physiotherapist available at least ≥ 18 hours a day.

#### Exclusion criteria for ICUs

- ICUs with structured multidisciplinary rounds, defined as meetings (DMRs) ≥ 3 times per week, during weekdays, conducted by a board-certified intensivist and documented in medical records with fixed visit length (>5 min / patient), using some supporting tool (checklist or standard form), goal-oriented, based on established protocols, including all the patients admitted to the ICU.
- ICUs with implemented monthly management of indicators (audit and feedback) with specific planning.
- Coronary / cardiac intensive care units or other specialized units (e.g, neurological, burned patients).
- Step-down or intermediate care units.
- ICUs without availability of renal replacement therapy.
- ICUs in which coordinators are board-certified intensivists and qualified as Master of Business Administration (or an equivalent).

#### Inclusion criteria for patients

- All consecutive patients admitted to the ICU, aged 18 years or older after the beginning of the trial.

#### Exclusion criteria for patients

- Admission to the ICU due to justice-related issues (since in such circumstances the ICU admission or discharge may be determined by the law rather than by medical reasons).
- Previously included in the Telescope 2 study (for the primary outcome analysis).

### Interventions

The trial comprises three interventions, delivered in combination, via telemedicine:

- DMRs conducted by board-certified intensive care physicians.
- Coordinated care by a multidisciplinary team, involving a team of nurses, physiotherapists, and clinical pharmacists.
- Management strategy focused on quality improvement and patient safety, delivered by intensivists also qualified as management specialists, focused on quality improvement and patient safety.

DMR with an intensivist is a discussion led by remote board-certified intensivists. These DMRs will occur from Monday through Friday at a predetermined time, either in the morning or afternoon, based on the ICU’s preference, in centers equipped with environmental microphone/high-fidelity sound system, webcam, and high-resolution camera with remote web control. During the intervention period, all patients admitted to the participating ICUs will be exposed to the multifaceted remote intervention. On weekends and national holidays, DMRs will not be conducted, but there will be in place an urgent communication channel with the attending clinician and data collection will proceed as usual, allowing patient follow-up. The urgent communication channel is also available during weekdays.

DMR will be performed with the aim to establish diagnostic hypotheses, identify active problems, and develop a therapeutic plan until the next DMR. Remote intensivists will provide evidence-based recommendations tailored to the local structural context. Standardized electronic forms, filled by the tele-ICU physicians, will guide the discussions and support daily patient monitoring.

Coordinated care provided by a multidisciplinary team will collaborate with the tele-ICU physician and the on-site team, providing real time protocolized interventions, and providing evidence-based recommendations tailored to the local context. The team will include board-certified intensive care nurses, board-certified intensive care respiratory and physical therapists, and clinical pharmacists, supporting therapeutic plan, implementing preventive strategies, and promoting early recognition of clinical deterioration.

The management strategy focused on quality improvement and patient safety, were performed by intensivists with formal training in healthcare management. These activities will be conducted, daily, from Monday to Friday (excluding national holidays) at predetermined times, based on performance data (care performance indicators consolidated in a dashboard, built with software Shiny App, RStudio, Inc, R Foundation for Statistical Computing, Vienna, Austria). Monthly, the Tele-ICU medical coordinators will remotely meet with the local hospital leadership to discuss strategies for improving patient outcomes, develop strategies to prevent patient deterioration, evaluate readmissions within 48 hours, identify potentially avoidable bed days, and assess local practices related to the prevention and control of healthcare associated infections, including bloodstream infections, urinary tract infections with or without catheter use, tracheitis, pneumonia, and ventilator associated pneumonia. This integrated management intervention aims to enhance healthcare value by ensuring quality, safety, and efficiency in patient care.

### Outcomes

#### Primary outcome

The primary outcome of this trial at the patient level is ICU length of stay (LOS) defined as the time interval in hours between patients’ ICU admission and the moment of ICU physical discharge times (i.e., transfer to another care facility or another hospital) or ICU death, as defined by the hospital’s system date and time. Date and time will be entered by the health care worker responsible for data collection. ICU LOS will be derived in 24 hours periods with decimal place. ^(11)^

#### Secondary outcomes

The secondary outcomes of this study include assessing the impact of interventions implemented through telemedicine compared with a control period in the following outcomes:

- ICU mortality, defined as death by any cause during the index ICU admission.
- In-hospital mortality, defined as death by any cause during the index hospital admission.
- Ventilator-free days during the first 28 days, defined as the number of days alive and free from mechanical ventilation for at least 24 consecutive hours. Patients who died before weaning were deemed to have zero ventilator-free days, and patients discharged from the hospital before 28 days were considered alive and free from mechanical ventilation at 28 days.
- ICU readmission within 48 hours. ^(12)^
- Early reintubation (<48h after elective extubation).
- Ventilator-associated events, defined as either an increase in the daily minimum positive end-expiratory pressure (PEEP) of ≥3 cmH_2_O sustained for at least two consecutive calendar days following a period of at least two days of stable or decreasing PEEP, or an increase in the fraction of inspired oxygen (FiO_2_) by ≥20 percentage points sustained for at least two consecutive days after a minimum of two days of stable or decreasing FiO_2_ levels. ^(13)^
- Accidental extubation rate.

#### Process-of-care and quality indicators

- Patient mobilization density in the ICU, defined as the number of days the patient has undergone any form of mobilization, including passive movement of upper or lower limbs in bed, sitting on the edge of the bed or in a chair, or walking. ^(14)^
- Adherence to maintaining the head-of-bed elevation (30°-45°).
- Adequate prevention of venous thromboembolism. ^(15)^
- Rate of patient-days under adequate sedation [defined by Richmond agitation and sedation scale (RASS) = −3 to +1]. ^(16)^
- Rate of patients-days with oral or enteral nutrition.
- Rate of patients with adequate glycemic control (defined as blood glucose = 70 to 180mg/dL). ^(9, 15)^
- Rate of patients-days within normoxemia (defined as peripheral oxygen saturation = 92 to 96%). ^(9)^
- Rate of central venous catheter use.
- Central venous catheter dwell time.
- Rate of indwelling urinary catheter use.
- Indwelling urinary catheter dwell time.

#### Unit-level organizational outcomes

- Classification of the unit according to the profiles defined by standard resource use (SRU) and standardized mortality rate (SMR). ^(17)^ SRU reflects the observed-to-expected rate of resource utilization, estimated as ICU LOS for surviving patients and adjusted for the patient’s severity of illness. SMR reflects the observed / expected rate, according to acute physiology score (SAPS 3) of hospital deaths. ^(17)^ The profiles are a combination of SMR (above or below median) and SRU (above or below median): Each unit can be assigned to one of four groups: ‘most efficient’ (SMR and SRU <median); ‘least efficient’ (SMR, SRU >median); ‘overachieving’ (low SMR, high SRU), ‘underachieving’ (high SMR, low SRU). ^(17)^

The follow-up period to define all outcomes will be truncated at 90 days while in the hospital from ICU admission.

### Participant timeline and follow-up

Eligible ICUs will be randomized and allocated to one of five sequences (Figure 1). Each sequence will include at least a 3-month control period, followed by a 3-month transition period, and at least a 3-month intervention period. The duration of intervention varies by sequence, ranging from 19 months in sequence one to 3 months in sequence five. The total study duration is 25 months. As a stepped-wedge, all ICUs will receive all interventions, but at different times. Patients will be followed up until hospital discharge.

**Figure 1.**
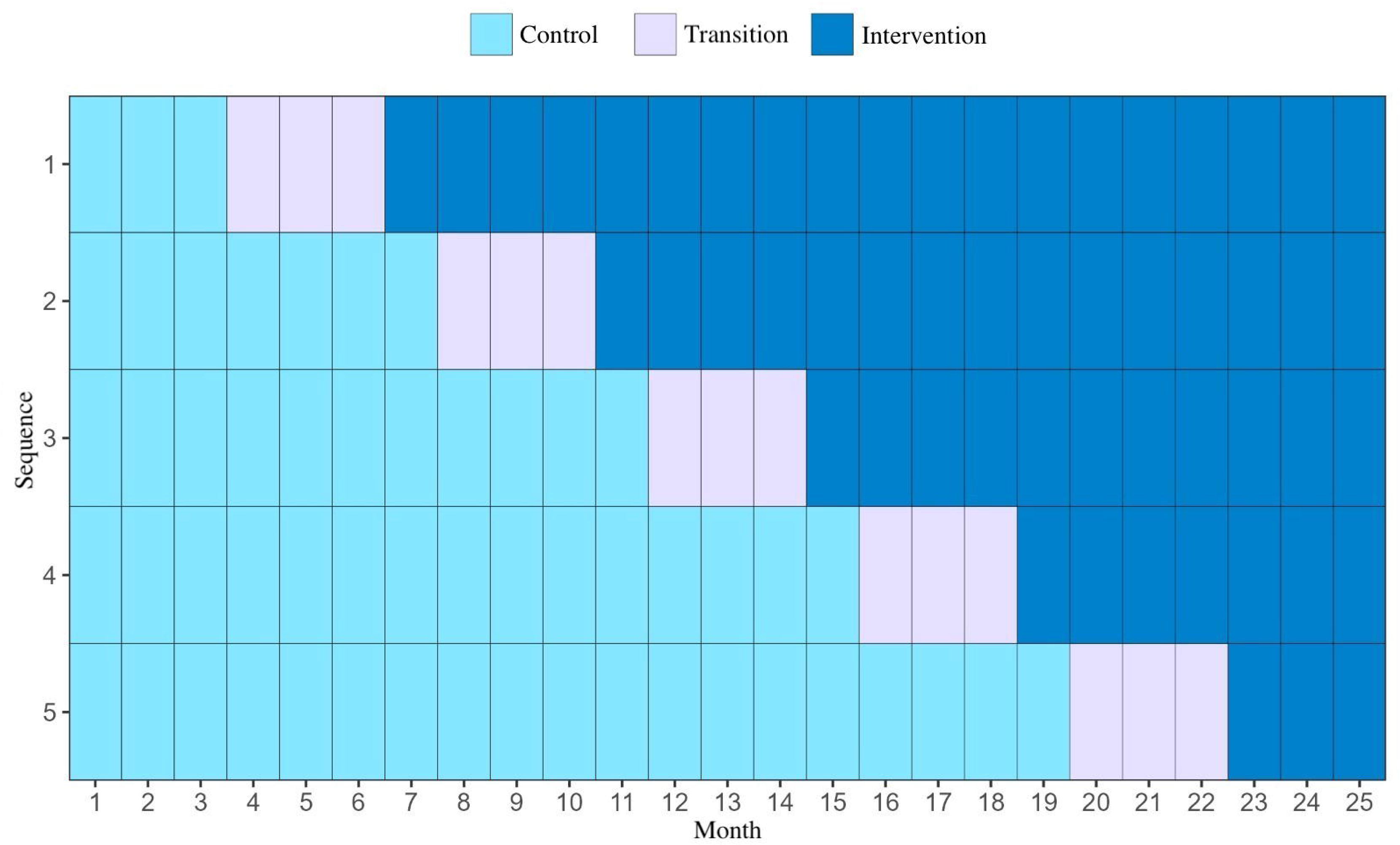
Schematic diagram illustrating the trial timeline, control period, randomization, transition period, intervention period, and follow-up.

Aiming maximum engagement, the study’s executive committee will hold in-person meetings with all centers before the study intervention. Those meetings will occur within 30 days of the center’s transition date, with the goal of reviewing the study’s key aspects and aligning on the proposed interventions. Additionally, aiming to maximize local teams’ engagement, remote tele-ICU professionals will always start intervention, primary *in loco*, transitioning to telemedicine afterwards.

### Calculation of sample size and recruitment

A total sample size of 18,750 to 25,000 patients will be required to detect a 1.1-day reduction in ICU length of stay compared to baseline, attributed to the intervention package, with a significance level of 5% and a minimum power of 95%. This variation in total sample size is due to different estimates of patients per period in the 25 Brazilian ICUs in question. It is estimated that there will be a variation of 30 to 40 patients recruited per month per ICU. This sample size was estimated based on simulations using data from the TELESCOPE 1 trial and it is specified elsewhere. ^(9)^

### Sequence generation, allocation and implementation

The 25 ICUs will be randomized into one of five sequences (Figure 1). The allocation sequence will be computer-generated by a statistician (TMS) and a clinical trialist from the study’s executive committee (OTR) using random numbers, stratified by region (south/southeast, and central-west/north/northeast) and baseline length of stay, estimated from the first two months of the study within the initial period. Length of stay will be categorized into two strata: below the regional median and above the regional median. All ICUs will be randomized simultaneously, with the ICU serving as the unit of randomization, since the intervention is applied to the entire multidisciplinary team. To ensure allocation concealment, only the statistician responsible for the randomization list will know the sequence and will reveal the next sequence 30 days before the start of the transition. This period is necessary to organize the in-person meeting with the center. The allocation concealment will be maintained until the end of the study.

### Blinding (masking)

The intervention is open, due to the nature of the study, i.e., DMRs, quality improvement meetings and delivery of evidence-based clinical protocols. Statisticians, and research team will be blinded for analysis, and discussion.

### Data collection

Forms detailing data collection during DMRs conducted by the physicians, nurses, physiotherapists, and clinical pharmacists are included in the Additional file 1. At the patient level, frequency of data collection is outlined in Table 2, and the following data will be collected:

**Table 2.** Patient data collection schedule.

#### At the time of ICU admission

- Identifier, date of birth, sex, race, main reason of ICU admission (adapted from Acute Physiology and Chronic Health disease Classification System-APACHE III), ^(18)^ readmission status.
- Anthropometric characteristics, comorbidities (adapted from SAPS 3), ^(19)^ functional status before hospitalization.
- Respiratory, cardiovascular and renal support. ^(9, 20, 21)^
- Diet, sedation status, and vasoactive drugs. ^(9)^
- Presence of devices: central venous catheter (CVC), arterial line, permanent catheters, urinary catheter, oro/nasotracheal catheter and tracheostomy.
- Date and time of hospital admission.
- Date and time of ICU admission.
- SAPS 3 score. ^(19)^
- Sequential organ failure assessment (SOFA) score. ^(22)^

#### Throughout the ICU admission

- Documented goals from the DMR (additional file 1).
- Documented discharge order status, defined as any mention to readiness to discharge or ICU transference order.
- Mechanical ventilation (MV) status and MV parameters.
- Peripheral saturation of hemoglobin measured by pulse oximetry range for patients on oxygen therapy.
- Head of bed elevation for patients under MV.
- Spontaneous respiratory test, accidental extubation or reintubation events.
- Need of vasoactive drugs and renal replacement therapy.
- Continuous sedative infusion and light sedation strategy (reduction/daily interruption).
- Daily value (categorized below, above or within −3 to +1 range) of the RASS for patients undergoing continuous sedation at a predetermined time. ^(16)^
- Glasgow coma scale.
- Adequacy of venous thromboembolism prophylaxis [considered adequate when patient is bedridden without any of the following exclusion criteria: active bleeding, stress gastric ulcer, uncontrolled arterial hypertension (>180/110 mm Hg), coagulation disorder, allergy, kidney failure (Cl <30 mL/min), ocular or cranial surgery in last 2 weeks, and lumbar puncture in last 24 hours)].
- Presence of oral or enteral nutrition.
- Glycemic control: considered adequate if between 70 and 180 mg/dL. ^(9)^
- Notification of healthcare-related infection episodes according to Centers for Disease Control and Prevention (CDC) criteria:

`o` Central line-associated bloodstream infection. ^(23)^
`o` Catheter-associated urinary tract infection. ^(24)^
- Mobilization activity.
- Date and time of CVC insertion for patients undergoing CVC insertion.
- Date and time of withdrawal of CVC for patients undergoing CVC insertion.
- Date and time of indwelling urinary catheter (IUC) insertion for patients submitted to IUC insertion.
- Date and time of withdrawal of IUC for patients undergoing IUC insertion.
- Documentation of decisions for limiting the life support considering any mention to withholding or withdrawing in the medical records.

#### At the time of ICU discharge

- Date and time of ICU discharge.
- ICU outcome: discharge to ward, hospital transfer, death.

#### At the time of hospital discharge

- Date and time of hospital discharge.
- Hospital outcome: hospital transfer, death.

### Data management

Trained healthcare workers will collect data, without any involvement from the study committees and investigators. A standard case report form was developed for the trial (additional file 2), with extensive validation and piloting aiming clarity and consistency.

Data will be entered using an electronic case report form in the Research Electronic Data Capture (REDCap®, USA) platform via internet, hosted on a server at the Einstein Hospital Israelita /São Paulo-Brazil. ^(25, 26)^ The system has different functionalities, including patient registration, data entry, data validation, data reports, data quality, data resolution workflow, audit trail, and data export for statistical analysis. ^(25, 26)^ Local investigators directly input data into the system, with comprehensive usage instructions always available to investigators. Electronic files will be securely stored at Einstein Hospital Israelita servers, in a controlled and confidential environment, with restricted access, following best practices. Regular remote data monitoring will promptly identify irregular patterns, inconsistencies, credibility concerns, or anomalies using predefined queries within the system. Missing or outlier data values will be individually reviewed, and follow-up reports are regularly reviewed by the coordinating center to ensure consistency and completeness. Continuous efforts will be made to complete or rectify data whenever possible.

### Database cleaning and locking

Database will be locked once all data have been entered and all discrepancies or missing data have been addressed. After this review, the database will be locked and prepared for statistical analysis. At this stage, access permissions for all investigators will be revoked, and the database will be archived.

### Statistical methods

All analyses will be thoroughly described in a statistical analysis plan (SAP), which will be concluded and submitted for publication elsewhere before the database is closed and analyses begin. Briefly, primary statistical analyses will be performed according to the intention-to-treat principle. All outcomes at the patient-level will be performed using models that account for correlated data within each ICU (i.e., ICU as a cluster) with generalized linear mixed models and adjusted by pre-specified covariates, as they will be specified in the SAP. Four subgroups were prespecified for analysis: type of admission (medical vs surgical), telemedicine experience (centers with vs. without prior telemedicine use), SAPS3 score (categorized by tertiles), mechanical ventilation status at admission (invasive vs. non-invasive x none), and ICU performance at baseline (four categories: most efficient, least efficient, overachieving, underachieving; each classification will be made based on the first three months).

All analyses will be performed using R software R (V.4.2.0, the version will be updated at the time of analysis).

### Auditing

The TELESCOPE 2 study is subjected to audit by Einstein Research Integrity Committee at any time, independently of the institutional review board, and the research team, following standard procedures.

## ETHICAL CONSIDERATIONS

The study will be performed according to national and international guidelines, adhering to the principles of the Declaration of Helsinki and the Act for Medical Research Involving Humans. The study was approved by local Research Ethics Committee of the coordinating study center (Einstein Hospital Israelita, CAAE: 69575123.0.1001.0071), as well as by local IRBs from each one of the 25 participant centers, in compliance with Brazilian legislation. A specialist in regulatory processes will oversee and support the local teams. Any modifications to the protocol that may affect the study’s development, potential benefits, or safety – including changes in the objectives, design, study population, sample size, interventions or relevant management aspects – require protocol amendments. These amendments should be submitted for approval to the IRB of the coordinating center and all the IRBs at participating centers.

## SAFETY AND MONITORING

### Adverse events and interim analyses

Considering the study intervention incorporates the best available evidence for the care of critically ill patients in ICUs, interventions will be decided by consensus, and always validated by *in loco* professionals, and no major inherent risks are anticipated in the trial’s execution, interim analyses are not planned. As a result, the constitution of a formal data monitoring committee was deemed unnecessary. While adverse events are not expected, they will be requested to be reported by local researchers, data assistants, and attending physicians.

### Patient information and informed consent

The need for patients’ written informed consent was waived in all 25 centers. ^(27–31)^ Consent was obtained at the cluster level, with the hospital director and the head of the ICU (physician) responsible for signing the consent form in some cases, when requested by local IRBs. ^(27–31)^ This approach was approved by all the local IRBs (including the coordinating center and the IRBs of the 25 participating ICUs).

### Data confidentiality

Patient and the participant centers will be identified by corresponding numbers in the electronic data collection form to maintain anonymity. Data obtained from medical records will be handled confidentially and stored keeping restricted access, only authorized to part of the research team (directly linked to data collection and data management). Anonymization of all data, both in provisional and final reports, will be ensured, with no identifiable information disclosed. Research sites must securely store all data for the duration specified by the study and in accordance with local regulations. After this period, data must be securely incinerated (if physical) and/or excluded (if digital) to prevent unauthorized access. Research team is committed to taking all necessary precautions to guarantee data confidentiality throughout the study and beyond.

## PUBLICATION AND ADMINISTRATIVE ASPECTS

### Coordinating center

The coordinating center for the study is Einstein Hospital Israelita in São Paulo, Brazil. Its responsibilities include planning and overseeing the study, preparing the study protocol and data collection forms, developing the operations manual, managing and ensuring data quality, conducting statistical analyses, and preparing the final manuscript.

### Public disclosure and publication policy

The Telescope 2 group will publish the study findings regardless of the results. The main manuscript will be submitted by the executive and writing committee on behalf of the Telescope 2 research group.

### Organization

Local Principal Investigators (PIs) will be responsible for site recruitment and overseeing the proper execution of the study. PIs at each participating center will provide both scientific and structural leadership, ensuring that all required local ethical and regulatory approvals are obtained before patient enrollment. They will also train and supervise local data assistants, ensuring the accuracy of data collection and proper entry of data into the electronic medical record. The Steering Committee (composed by invited and highly experienced Brazilian researchers in the critical care field) provides high-level, independent oversight, ensuring patient safety, scientific integrity, and adherence to protocols, while the Executive Committee (EC) / Trial Management Group makes critical, executive decisions on the study’s direction, resource allocation, and routine problem-solving.

## TRIAL STATUS

This manuscript describes the protocol for the TELESCOPE 2 trial (original version 1, approved on June 28, 2023). The baseline period started on January 8, 2024, followed by randomization on March 8, 2024. The first transition period started on April 8, 2024, with the first interventions starting on July 8, 2024. At the time of the manuscript’s submission, data collection for the trial was ongoing and is expected to be completed by January 31, 2026.

## Data Availability

All data produced in the present study are available upon reasonable request to the authors

## Acknowledgements

The authors would like to thank the central TELESCOPE 2 team, data collection team of each participating ICU, as well as the Einstein Hospital Israelita, and the Brazilian Ministry of Health.

## COLLABORATORS

### Coordinating Center

Einstein Hospital Israelita

### Executive and steering committee

Renato Carneiro de Freitas Chaves, Bruna Gomes Barbeiro, Maura Cristina dos Santos, Tiago Mendonça dos Santos, Thiago Domingos Corrêa, Otavio T. Ranzani, Adriano José Pereira.

### Advisory and scientific committee

Alexandre Biasi Cavalcanti, Ary Serpa Neto, Carlos Henrique Sartorato Pedrotti, Fernando Zampieri, Guilherme de Paula Pinto Schettino, Jorge Ibrain Figueira Salluh, Leandro Utino Taniguchi, Leonardo José Rolim Ferraz, Luciano Cesar Azevedo, Otávio Berwanger, Regis Goulart Rosa, Renata Albaladejo Morbeck, Rodrigo Biondi, Suzana Margareth Lobo.

### Experts Committee (multidisciplinar team)

Alessandra Gomes Chauvin, Aline Cristina Pedroso, Barbara Barduchi, Beatriz Rocha Monteiro, Camila de Carvalho Gambin, Cilene Saghabi, Eliton Paulo Leite Lourenço, Erika Yumiko Kumoto, Fabiana Rossi Varallo, Fernanda Paulino Fernandes, Flavia Oliveira Rodrigues, Flavia Veronezi Stankevicius, Gabrielli Pare Guglielmi, Gean Carlos Alves Moraes, Giovana Roberta Zelezoglo, Lidiane Soares Sodre da Costa, Luciana Laversveiler Moraes da Costa, Jessica Tamiris Romano, Joao Paulo Victorino, Marcele Pessavento, Raquel Afonso Caserta Eid, Renata de Souza Cyrino, Roberta Gonsalez dos Santos, Silvana Maria de Almeida, Tatiana Aporta Marins.

### Experts Committee (management team)

Amanda Valle, Ana Cláudia Ferraz, Ana Lúcia Martins da Silva, Bruno de Arruda Bravim, Bruno Mazza, Daiane Seger, Felipe Maia de Toledo Piza, Gilberto Friedman, Glauco Whestphal, Guilherme de Paula Pinto Schettino, Gustavo Faissol Janot de Matos, Haggéas da Silveira Fernandes, Hipólito Carraro Jr., Joan Castro, Juliana Anacleto, Marcele Pesavento, Murillo Santucci Cesar de Assunção, Nelson Akamine, Niklas Soderberg Campos, Walace de Souza Pimentel.

### Research back office

Ana Cristina Lagoeiro Patrocinio da Cruz, Andrea de Carvalho, Lucelio De Sousa Rocha, Lenine Melo Lino, Rodrigo Flor de Moura.

### Funding

The Brazilian Ministry of Health (Institutional Development Program of the Unified Health System-PROADI SUS) was the primary source of funding, including costs of physician services, purchase of equipment (hardware) for Telemedicine sessions, hiring of local professionals for data collection and travel expenses for training and monitoring. The same funding also covered costs related to the regulatory part of the study-data collection, monitoring, data curation and statistical support. The Einstein Hospital Israelita allocated time of professionals and specialists who sat on the executive and steering committee of the study, as well as assign its Telemedicine service system.

## LIST OF ABBREVIATIONS

APACHE III: Acute Physiology and Chronic Health disease Classification System.
CDC: Centers for Disease Control and Prevention.
CVC: central venous catheter.
DMR: daily multidisciplinary rounds.
FiO_2_: fraction of inspired oxygen.
ICU: intensive care unit.
IUC: indwelling urinary catheter.
LOS: length of stay.
MV: mechanical ventilation.
PEEP: positive end expiratory pressure.
SAPS: Simplified acute physiology.
SOFA: Sequential organ failure assessment.
SMR: standardized mortality rate.
SRU: standard resource use.

## LIST OF HOSPITALS PARTICIPATING IN THE STUDY AND LOCAL PRINCIPAL INVESTIGATORS (LPI)

- Hospital Ana Nery, Salvador, BA. LPI: Luiz Carlos Santana Passos.
- Hospital das Clínicas Luzia de Pinho Melo, Mogi das Cruzes, SP. LPI: Daniella de Rezende Duarte Maksymczuk.
- Hospital de Emergência Dr. Daniel Houly, Arapiraca, AL. LPI: Guacyra Almeida.
- Hospital de Emergência e Trauma Dom Luiz Gonzaga Fernandes, Campina Grande, PB. LPI: Dagjane Martins Frazão
- Hospital Dr. Genesio Rego, São Luís, MA. LPI: Thamyris Danusa da Silva Lucena.
- Hospital Dr. Leo Orsi Bernardes, Itapetininga, SP. LPI: Vivian Menezes Irineu.
- Hospital Mangabeira Governador Tarcísio Burity, João Pessoa, PB. LPI: Madson Mariz Melo Tavares.
- Hospital Estadual de Doenças Tropicais Dr. Anuar Auad, Goiânia, GO. LPI: Adriana Rocha de Carvalho Sousa.
- Hospital Estadual de Luziânia, Luziânia, GO. LPI: Renata Meireles Roriz de Moraes, Patricia Alves de Castro Porto Marinho.
- Hospital Geral de Roraima, Boa Vista, RR. LPI: Mauro Shosuka Asato,
- Hospital Maternidade e Pronto Socorro Santa Lúcia, Poços de Caldas, MG. LPI: Ricardo Reinaldo Bergo.
- Hospital Municipal de Santarém, Santarém, PA. LPI: Antonio Carlos Alves da Silva.
- Hospital Municipal Padre Germano Lauck, Foz do Iguaçu, PR. LPI: Roberto de Almeida.
- Hospital Municipal Salgado Filho, Rio de Janeiro, RJ. LPI: Claudia Valeria Pereira de Oliveira.
- Hospital Municipal Senhora Santana, Brasília de Minas, MG. LPI: Breno Mendes Cardoso.
- Hospital Nossa Senhora das Dores, Itabira, MG. LPI: Thállassa Fürst Guerra Procópio, Edilene Aparecida dos Santos.
- Hospital Padre Máximo, Venda Nova do Imigrante, ES. LPI: Monica Paula de Deus Cezati, Maria Caroline de Souza Marques.
- Hospital Regional de Barbacena, Barbacena, MG. LPI: Paulo Roberto Rezende.
- Hospital Regional De Cáceres Dr. Antônio Fontes, Cáceres, MT. LPI: Thayla de Arruda Moura, Dennislaine Alves Lima Dantas.
- Hospital Regional Justino Luz, Picos, PI. LPI: Raimundo de Carvalho Reis Neto.
- Hospital Região Leste (Paranoá), Brasília, DF. LPI: Sidney Sotero.
- Hospital São Lucas, Garça, SP. LPI: Barbara Cristine Teixeira Ferreira.
- Hospital Tramandaí, Tramandaí, RS. LPI: Jocelaine Lencina, Juliana da Silva Silveira.
- Irmandade da Santa Casa de Misericórdia de Mococa, Mococa, SP. LPI: Luiz Otavio da Silva, Camila Gabriel Carraro.
- Santa Casa de Paranavaí, Paranavaí, PR. LPI: Bruno Cesar Pereira Leal.

## Notes

### Competing Interest Statement

The authors have declared no competing interest.

### Clinical Trial

Clinicaltrials NCT05960994, Brazilian Registry of Clinical Trials RBR-342wxn9, and Universal Trial Number U1111-1298-9799.

### Author Declarations

The study was approved by local Research Ethics Committee of the coordinating study center (Einstein Hospital Israelita, CAAE: 69575123.0.1001.0071), as well as by local IRBs from each one of the 25 participant centers, in compliance with Brazilian legislation.

