## Additional file 1 for "Evaluation of the Clinical Impact of Different Telemedicine Practices in Intensive Care Units: a Stepped-wedge Cluster Randomized Clinical Trial (TELESCOPE 2): study protocol"

**Authors:** Adriano José Pereira, Bruna Gomes Barbeiro, Tiago Mendonça dos Santos, Maura Cristina dos Santos, Thiago Domingos Corrêa, Alexandre Biasi Cavalcanti, Ary Serpa Neto, Carlos Henrique Sartorato Pedrotti, Fernando Zampieri, Guilherme de Paula Pinto Schettino, Jorge Ibrain Figueira Salluh, Leandro Utino Taniguchi, Leonardo José Rolim Ferraz, Luciano Cesar Azevedo, Otávio Berwanger, Regis Goulart Rosa, Renata Albaladejo Morbeck, Rodrigo Biondi, Suzana Margareth Lobo, Renato Carneiro de Freitas Chaves, Otavio Ranzani.

**Additional file 1 contains the following document:**

1. Form to support the conduction of daily multidisciplinary rounds (DMR) by physicians………………………………………………………………………….**3**
2. Form to support the conduction of DMR by nurses………………………...…**9**
3. Form to support the conduction of DMR by physiotherapists……………....**11**
4. Form to support the conduction of DMR by clinical pharmacists…………..**13**

**Daily multidisciplinary rounds - physician**

**Full name of the patient:__________________________________________**

**Social name of the patient:________________________________________**

**Individual Taxpayer Registry (CPF):_________________________________**

**Gender:** Female🞎	Male 🞎

**Medical record number:___________________________________________**

**Ward:** 🞎 ICU

**Date of birth:____________________________________________________**

**Patient's age (auto-filled):_________________________________________**

**Weight:_________________________________________________________**

**Height**:**_________________________________________________________**

**Hospital admission date:__________________________________________**

**Days hospitalized (auto-filled):_____________________________________**

**ICU admission date:______________________________________________**

**Days in ICU (auto-filled):__________________________________________**

**Admission diagnosis:_____________________________________________**

**Comorbidities**

| 🞎 Diabetes Mellitus | 🞎 Hypertension |
| --- | --- |
| 🞎 Chronic Obstructive Pulmonary Disease | 🞎 Asthma |
| 🞎 Chronic Heart Disease | 🞎 Acute Myocardial Infarction |
| 🞎 Peripheral vascular insufficiency | 🞎 Cirrhosis |
| 🞎 Stroke | 🞎 Congestive Heart Failure |
| 🞎 Psychiatric disorders | 🞎 Dementia |
| 🞎 Seizures | 🞎 Chronic Kidney Disease |
| 🞎 Post-transplant | 🞎 Cancer |
| 🞎 Not applicable | 🞎 Other |

**Allergies:** 🞎 Yes	🞎 No

**Patient's medications (before hospital admission)**

| 🞎 Antiarrhythmic | 🞎 Anticoagulant |
| --- | --- |
| 🞎 Vasodilator | 🞎 Diuretic |
| 🞎 Oral hypoglycemic | 🞎 Insulin |
| 🞎 Anxiolytic | 🞎 Antidepressant |
| 🞎 Other | 🞎 Not applicable |

**Previous functionality:**

🞎 Independent	🞎 Requires assistance	🞎 Restricted/Bedridden

**Watcher patient:** 🞎 Yes	🞎 No

**Form type:** 🞎 Tele-round	🞎 Tele-visit

**Hospitalization Summary**

**Admission:______________________________________________________**

**Relevant Findings:________________________________________________**

**Active Problems:_________________________________________________**

**Name of Physician:________________________________________________**

**NEUROLOGICAL SYSTEM**

**Glasgow Coma Scale:**

🞎 15	🞎 13 to 14	🞎 10 to 12	🞎 6 to 9	🞎 <6

**Sedation:** 🞎 Yes	🞎 No

**Pain:** 🞎 Yes	🞎 No

**Delirium**: 🞎 Yes	🞎 No

**HEMODYNAMIC SYSTEM**

**Laboratory Tests**

| pH**:______________________** | Arterial oxygen saturation (SaO_2_)**:_______** |
| --- | --- |
| Partial pressure of oxygen (PO_2_)**:_** | Venous oxygen saturation (SvO_2_)**:_______** |
| Arterial partial pressure of carbon dioxide (PaCO_2_)**:_____________** | Lactate**:___________________________** |
| Venous partial pressure of carbon dioxide (PvCO_2_)**:______** | Hemoglobin**:________________________** |
| Bicarbonate (HCO_3_)**:___________** | GAP PCO2**:_______________________** |
| Base excess (BE)**:_____________** |  |

**Vasopressor Use (e.g., norepinephrine, vasopressin, adrenaline):** 🞎 Yes	🞎 No

**Inotropic Use (e.g., dobutamine, milrinone, levosimendan):** 🞎 Yes	🞎 No

**Vasodilator use (e.g., nitroglycerin, nitroprusside):** 🞎 Yes	🞎 No

**Mean arterial pressure target:**

Minimum:**_______________________________________________________**

Maximum:**_______________________________________________________**

**Antiarrhythmic use:** 🞎 Yes	🞎 No

**SOFA cardiovascular:**

🞎 No hypotension

🞎 MAP < 70 mmHg

🞎 Dopamine > 5 or any dose of dobutamine

🞎 Dopamine >15 or norepinephrine/adrenaline > 0.01

🞎Norepinephrine/adrenaline > 0.1

**RESPIRATORY SYSTEM**

**Laboratory tests**

| Fraction of Inspired Oxygen (FiO_2_)**:** | pH**:_______________________________** |
| --- | --- |
| Partial pressure of oxygen (PO_2_)**:__** | Arterial partial pressure of carbon dioxide (PaCO_2_)**:_** |
| Bicarbonate (HCO_3_)**:____________** | Base excess (BE)**:_______________________** |
| Arterial oxygen saturation (SaO_2_)**:_** | PaO2 / FiO2 ratio**:_________________________** |

**Respiratory support:**

🞎 Ambient air

🞎 Oxygen mask or nasal cannula

🞎 Non-invasive ventilation

🞎 High-flow nasal cannula

🞎 Invasive mechanical ventilation

**Protective ventilation (at the time of visit):**

🞎 Tidal volume ≤ 6 ml/kg predicted weight

🞎 Peak pressure < 30 cmH_2_O

🞎 Driving pressure < 15 cmH_2_O

🞎 PEEP (ARDSNet table)

**Eligible for spontaneous breathing trial:** 🞎 Yes	🞎 No

**SOFA respiratory:**

🞎 PaO2 / FiO2 ratio ≥ 400

🞎 PaO2 / FiO2 ratio 300-399

🞎 PaO2 / FiO2 ratio 200-299

🞎 PaO2 / FiO2 ratio 100-199 + supplemental ventilation

🞎 PaO2 / FiO2 ratio <100 + supplemental ventilation

**NUTRITIONAL SUPPORT AND GLYCEMIC CONTROL**

**Laboratory tests**

| Total bilirubin**:_______________** | Direct bilirubin**:____________________** |
| --- | --- |
| Indirect bilirubin**:_____________** | alanine transaminase (ALT)**:_________** |
| aspartate transaminase (AST)**:__** | gamma-glutamyl transferase (GGT):**___** |
| Lipase**:____________________** | Amylase**:_________________________** |
| Alkaline Phosphatase (FA)**:____** | Albumin**:_________________________** |

**Is the patient being fed?** 🞎 Yes	🞎 No

**Bowel Movements:** 🞎 Present	🞎 Absent

**Did the patient have two or more blood glucose readings >180 mg/dl in 24 hours?** 🞎 Yes	🞎 No

**Any glycemic controls < 60 mg/dl?** 🞎 Yes	🞎 No

**SOFA hepatic:**

🞎 Total Bilirubin <1.2

🞎 Total Bilirubin 1.2 - 1.9

🞎 Total Bilirubin 2.0 - 5.9

🞎 Total Bilirubin 6.0 - 11.9

🞎 Total Bilirubin ≥12

**FLUID BALANCE, ELECTROLYTES, AND RENAL FUNCTION**

**Laboratory tests**

| pH**:_________________________** | Bicarbonate (HCO_3_)**:_______________** |
| --- | --- |
| Base excess (BE)**:_____________** | Potassium**:_______________________** |
| Sodium**:____________________** | Ureia**:___________________________** |
| Creatinine**:__________________** | Ionic calcium**:_____________________** |
| Chloride**:____________________** | Magnesium**:______________________** |
| Lactate**:_____________________** |  |

**Electrolyte imbalance:** 🞎 Yes	🞎 No

**On dialysis:** 🞎 Yes	🞎 No

**SOFA renal:**

🞎 Creatinine < 1.2

🞎 Creatinine 1.2 - 1.9

🞎 Creatinine 2.0 - 3.4

🞎 Creatinine 3.5 - 4.9 or urine output ≤ 500ml/24h

🞎 Creatinine ≥ 5 or urine output ≤ 200ml/24h

**HEMATOLOGY AND INFECTION**

**Laboratory tests**

| Hemoglobin**:___________________** | Hematocrit**:_____________________** |
| --- | --- |
| Leukocytes**:____________________** | Platelets**:_______________________** |
| Prothrombin Time**:_______________** | Partial Thromboplastin Time**:_______** |
| International normalised ratio (INR)**:_** | C-reactive Protein**:_______________** |

**Antibiotic Therapy:** 🞎 Therapeutic	🞎 Prophylactic	🞎 No antibiotics

**Infection Present:** 🞎 Yes	🞎 No

**SOFA hematology (platelet count):**

🞎 ≥ 150

🞎 100 - 149

🞎 50 - 99

🞎 20 - 49

🞎 < 20

**CLINICAL PHARMACOLOGY**

**Were drugs adjusted for renal function?** 🞎 Yes	🞎 No	🞎 Not applicable

**Medication Reconciliation:** 🞎 Total	🞎 Partial	🞎 Not applicable

**DEVICES AND PROCEDURES**

**Indwelling Urinary Catheter:** 🞎 Yes	🞎 No

**Central Venous Catheter:** 🞎 Yes	🞎 No

**Arterial Catheter:** 🞎 Yes	🞎 No

**Drains (chest/abdomen/subcutaneous/other):** 🞎 Yes	🞎 No

**PROPHYLAXIS**

**Is gastric prophylaxis indicated?** 🞎 Yes	🞎 No

**In use of gastric prophylaxis**? 🞎 Yes	🞎 No

**Is venous thromboembolism prophylaxis indicated?** 🞎 Yes	🞎 No

**In use of venous thromboembolism prophylaxis**	🞎 Yes	🞎 No

**MOBILIZATION**

**Can the patient be mobilized?** 🞎 Yes	🞎 No

**SKIN CONDITION**

**Skin Intact:** 🞎 Yes	🞎 No

**SUPPORT AND CONFLICT MANAGEMENT**

**Therapeutic Limitation:** 🞎 Yes	🞎 No

**PATIENT FLOW**

**Is the patient ready for discharge?** 🞎 Yes	🞎 No

**SOFA score (auto-filled):**___________________________

**Therapeutic Plan:________________________________________________**

_______________________________________________________________

_______________________________________________________________

_______________________________________________________________

**Daily multidisciplinary rounds - nursing**

**Full name of the patient:__________________________________________**

**Social name of the patient:________________________________________**

**Individual Taxpayer Registry (CPF):_________________________________**

**Gender:** Female🞎	Male 🞎

**Medical record number:___________________________________________**

**Ward:** 🞎 ICU

**Date of birth:____________________________________________________**

**Patient's age (auto-filled):_________________________________________**

**Weight:_________________________________________________________**

**Height**:**_________________________________________________________**

**Hospital admission date:__________________________________________**

**Days hospitalized (auto-filled):_____________________________________**

**ICU admission date:______________________________________________**

**Days in ICU (auto-filled):__________________________________________**

**Admission diagnosis:_____________________________________________**

**Comorbidities**

| 🞎 Diabetes Mellitus | 🞎 Hypertension |
| --- | --- |
| 🞎 Chronic Obstructive Pulmonary Disease | 🞎 Asthma |
| 🞎 Chronic Heart Disease | 🞎 Acute Myocardial Infarction |
| 🞎 Peripheral vascular insufficiency | 🞎 Cirrhosis |
| 🞎 Stroke | 🞎 Congestive Heart Failure |
| 🞎 Psychiatric disorders | 🞎 Dementia |
| 🞎 Seizures | 🞎 Chronic Kidney Disease |
| 🞎 Post-transplant | 🞎 Cancer |
| 🞎 Not applicable | 🞎 Other |

**Allergies:** 🞎 Yes	🞎 No

**Patient's medications (before hospital admission)**

| 🞎 Antiarrhythmic | 🞎 Anticoagulant |
| --- | --- |
| 🞎 Vasodilator | 🞎 Diuretic |
| 🞎 Oral hypoglycemic | 🞎 Insulin |
| 🞎 Anxiolytic | 🞎 Antidepressant |
| 🞎 Other | 🞎 Not applicable |

**Previous functionality:**

🞎 Independent	🞎 Requires assistance	🞎 Restricted/Bedridden

**Watcher patient:** 🞎 Yes	🞎 No

**Form type:** 🞎 Tele-round	🞎 Tele-visit

**Shift handover notes:_____________________________________________ ______________________________________________________________________________________________________________________________**

**Daily multidisciplinary rounds - physiotherapy**

**Full name of the patient:__________________________________________**

**Social name of the patient:________________________________________**

**Individual Taxpayer Registry (CPF):_________________________________**

**Gender:** Female🞎	Male 🞎

**Medical record number:___________________________________________**

**Ward:** 🞎 ICU

**Date of birth:____________________________________________________**

**Patient's age (auto-filled):_________________________________________**

**Weight:_________________________________________________________**

**Height**:**_________________________________________________________**

**Hospital admission date:__________________________________________**

**Days hospitalized (auto-filled):_____________________________________**

**ICU admission date:______________________________________________**

**Days in ICU (auto-filled):__________________________________________**

**Admission diagnosis:_____________________________________________**

**Comorbidities**

| 🞎 Diabetes Mellitus | 🞎 Hypertension |
| --- | --- |
| 🞎 Chronic Obstructive Pulmonary Disease | 🞎 Asthma |
| 🞎 Chronic Heart Disease | 🞎 Acute Myocardial Infarction |
| 🞎 Peripheral vascular insufficiency | 🞎 Cirrhosis |
| 🞎 Stroke | 🞎 Congestive Heart Failure |
| 🞎 Psychiatric disorders | 🞎 Dementia |
| 🞎 Seizures | 🞎 Chronic Kidney Disease |
| 🞎 Post-transplant | 🞎 Cancer |
| 🞎 Not applicable | 🞎 Other |

**Allergies:** 🞎 Yes	🞎 No

**Patient's medications (before hospital admission)**

| 🞎 Antiarrhythmic | 🞎 Anticoagulant |
| --- | --- |
| 🞎 Vasodilator | 🞎 Diuretic |
| 🞎 Oral hypoglycemic | 🞎 Insulin |
| 🞎 Anxiolytic | 🞎 Antidepressant |
| 🞎 Other | 🞎 Not applicable |

**Previous functionality:**

🞎 Independent	🞎 Requires assistance	🞎 Restricted/Bedridden

**Watcher patient:** 🞎 Yes	🞎 No

**Form type:** 🞎 Tele-round	🞎 Tele-visit

**Daily multidisciplinary rounds - clinical pharmacy**

**Full name of the patient:__________________________________________**

**Social name of the patient:________________________________________**

**Individual Taxpayer Registry (CPF):_________________________________**

**Gender:** Female🞎	Male 🞎

**Medical record number:___________________________________________**

**Ward:** 🞎 ICU

**Date of birth:____________________________________________________**

**Patient's age (auto-filled):_________________________________________**

**Weight:_________________________________________________________**

**Height**:**_________________________________________________________**

**Hospital admission date:__________________________________________**

**Days hospitalized (auto-filled):_____________________________________**

**ICU admission date:______________________________________________**

**Days in ICU (auto-filled):__________________________________________**

**Admission diagnosis:_____________________________________________**

**Comorbidities**

| 🞎 Diabetes Mellitus | 🞎 Hypertension |
| --- | --- |
| 🞎 Chronic Obstructive Pulmonary Disease | 🞎 Asthma |
| 🞎 Chronic Heart Disease | 🞎 Acute Myocardial Infarction |
| 🞎 Peripheral vascular insufficiency | 🞎 Cirrhosis |
| 🞎 Stroke | 🞎 Congestive Heart Failure |
| 🞎 Psychiatric disorders | 🞎 Dementia |
| 🞎 Seizures | 🞎 Chronic Kidney Disease |
| 🞎 Post-transplant | 🞎 Cancer |
| 🞎 Not applicable | 🞎 Other |

**Allergies:** 🞎 Yes	🞎 No

**Patient's medications (before hospital admission)**

| 🞎 Antiarrhythmic | 🞎 Anticoagulant |
| --- | --- |
| 🞎 Vasodilator | 🞎 Diuretic |
| 🞎 Oral hypoglycemic | 🞎 Insulin |
| 🞎 Anxiolytic | 🞎 Antidepressant |
| 🞎 Other | 🞎 Not applicable |

**Previous functionality:**

🞎 Independent	🞎 Requires assistance	🞎 Restricted/Bedridden

**Watcher patient:** 🞎 Yes	🞎 No

**Form type:** 🞎 Tele-round	🞎 Tele-visit
