## Additional file 1 for "Evaluation of the Clinical Impact of Different Telemedicine Practices in Intensive Care Units: a Stepped-wedge Cluster Randomized Clinical Trial (TELESCOPE 2): study protocol"

**Additional file 2**

**Authors:** Adriano José Pereira, Bruna Gomes Barbeiro, Tiago Mendonça dos Santos, Maura Cristina dos Santos, Thiago Domingos Corrêa, Alexandre Biasi Cavalcanti, Ary Serpa Neto, Carlos Henrique Sartorato Pedrotti, Fernando Zampieri, Guilherme de Paula Pinto Schettino, Jorge Ibrain Figueira Salluh, Leandro Utino Taniguchi, Leonardo José Rolim Ferraz, Luciano Cesar Azevedo, Otávio Berwanger, Regis Goulart Rosa, Renata Albaladejo Morbeck, Rodrigo Biondi, Suzana Margareth Lobo, Renato Carneiro de Freitas Chaves, Otavio Ranzani.

**Additional file 2 contains the following document:**

1. A standard case report form for the trial, used to collect data from all patients in the study through the Research Electronic Data Capture (REDCap®) system. Note: the data were gathered by trained healthcare workers without involvement from the study committees or investigators.

**CASE REPORT FORM**

**ELIGIBILITY CRITERIA**

**Patient is an adult at the time of admission (≥ 18 years)**

🞎 No 🞎 Yes

**Patient admitted to the ICU for medical reasons**

🞎 No (social, legal, or safety reasons) 🞎 Yes

**Patient admitted to the ICU through**

🞎 Unified Public Health System 🞎 Private health insurance

**PATIENT IDENTIFICATION**

**Full name of the patient:__________________________________________**

**Date of birth:____________________________________________________**

**Medical record number:___________________________________________**

**Individual Taxpayer Registry (CPF):_________________________________**

**Mother's full name:_______________________________________________**

**DEMOGRAPHY**

**Date and time of hospital admission:_____________________________**

**Sex:**

🞎 Female	🞎 Male

**Race:**

🞎 White

🞎 Black

🞎 Brown

🞎 Asian

🞎 Indigenous

🞎 Foreign

**Functional Capacity (consider the last 7 days prior to hospital admission):** 🞎 Independent for activities

🞎 Requires assistance

🞎 Bedridden/Restricted to bed

**ICU ADMISSION**

**Date and time of ICU admission:____________________________________**

**Age at ICU admission (auto-filled):__________________________________**

**Weight (Kg):_____________________________________________________**

**Height (cm):_____________________________________________________**

**Predicted body weight: (auto-filled):_________________________________**

**Reason for ICU admission**

🞎 Non-surgical

🞎 Surgical

**Specify the non-surgical reason for ICU admission:**

🞎 Cardiovascular/Vascular

🞎 Respiratory

🞎 Metabolic

🞎 Gastrointestinal

🞎 Neurological

🞎 Renal

🞎 Trauma

🞎 Hematological

🞎 Other

**Cardiovascular/Vascular**

🞎 Cardiogenic shock

🞎 Hypertension

🞎 Acute myocardial infarction

🞎 Decompensated heart failure

🞎 Cardiac arrest

🞎 Other

**Respiratory**

🞎 Asthma

🞎 Chronic obstructive pulmonary disease

🞎 Pneumonia

🞎 Pulmonary embolism

🞎 Other

**Metabolic**

🞎 Diabetic

🞎 ketoacidosis

🞎 Metabolic coma

🞎 Drug overdose

🞎 Other

**Gastrointestinal**

🞎 Liver failure

🞎 Gastrointestinal bleeding

🞎 Other

**Neurological**

🞎 Stroke

🞎 Seizures

🞎 Neuromuscular disease

🞎 Subarachnoid hemorrhage

🞎 Meningitis

🞎 Other

**Renal**

🞎 Urinary tract infection

🞎 Other

**Trauma**

🞎 Isolated head trauma or polytrauma

🞎 Polytrauma without head trauma

**Hematological**

🞎 Coagulopathy / Neutropenia / Thrombocytopenia

🞎 Other

**Specify the surgical reason for ICU admission**

🞎 Cardiovascular/Vascular

🞎 Respiratory

🞎 Gastrointestinal

🞎 Neurological

🞎 Trauma

🞎 Orthopedic

🞎 Gynecological

🞎 Renal/Urological

**Cardiovascular/Vascular**

🞎 Aortic surgery

🞎 Valve surgery

🞎 Peripheral vascular disease

🞎 Carotid endarterectomy

🞎 Coronary artery bypass graft

🞎 Other

**Respiratory**

🞎 Thoracic cancer surgery

🞎 Empyema drainage

🞎 Other

**Gastrointestinal**

🞎 Inflammatory abdomen (e.g., appendicitis, cholecystitis)

🞎 Obstructive abdomen

🞎 Perforated abdomen

🞎 Intestinal cancer

🞎 Gastrointestinal bleeding

🞎 Other

**Neurological**

🞎 Hemorrhagic stroke

🞎 Craniotomy for neoplasia

🞎 Spinal surgery

🞎 Sub/epidural hematoma drainage

🞎 Subarachnoid hemorrhage

🞎 Other

**Trauma**

🞎 Isolated head trauma or polytrauma

🞎 Polytrauma without head trauma

**RESOURCE UTILIZATION AND SUPPORT THERAPY IN THE FIRST HOUR**

**Ventilatory support**

🞎 Invasive (e.g., endotracheal intubation, tracheostomy)

🞎 Non-invasive (e.g., BiPAP, CPAP, high-flow nasal cannula)

🞎 None

**Vasoactive drug use** (e.g., adrenaline, dobutamine, dopamine, noradrenaline, vasopressin)

🞎 No 🞎 Yes

**Dialysis** (dialysis performed within the first hour of admission)

🞎 No 🞎 Yes

**Sedation**

🞎 No 🞎 Yes

**Richmond Agitation-Sedation Scale (RASS):_________________________**

**Glasgow Coma Scale:____________________________________________**

**SAPS 3:________________________________________________________**

**Sofa score 3 (**within the first 24 hour of admission)**:______________________**

**CLINICAL DAILY COLLECTION FORM**

**Evaluation Date:______________________**

**Is the patient still physically admitted to the ICU?**

🞎 No. The patient was discharged to a ward or transferred to another ICU within the same hospital or a different institution.

🞎 No. The patient temporarily left for an exam at another institution and returned within 24 hours.

🞎 No. The patient temporarily left for an exam at another institution and returned after more than 24 hours.

🞎 Yes. The patient remains admitted to the ICU.

**Are the objectives and conduct from the visit recorded in the medical chart for the care team to follow?**

🞎 No 🞎 Yes

**Has discharge been requested or prescribed, but the patient remains in the ICU?**

🞎 No 🞎 Yes

**Has the patient been under invasive mechanical ventilation in the last 24 hours?**

🞎 No 🞎 Yes

**Mechanical Ventilation Status**

🞎 Orotracheal intubation

🞎 Endotracheal extubation

🞎 New orotracheal intubation

🞎 Tracheostomy

**Maximum PEEP value:______________________**

**Maximum FiO2 value:______________________**

**Is the ventilatory mode controlled/assisted?**

🞎 No 🞎 Yes

**Currently tidal volume (ml):______________________**

**What is the patient’s RASS (Richmond Agitation-Sedation Scale) score at this moment?___________________________________________________**

**Is there a record of a spontaneous breathing test?**

🞎 No 🞎 Yes

**Was there an accidental extubation?**

🞎 No 🞎 Yes

**Was the patient extubated and re-intubated within the last 48 hours?**

🞎 No 🞎 Yes

**Is the head of the bed currently elevated to 30º or more?**

🞎 No 🞎 Yes

**Has the patient experienced a temperature below 36ºC or above 38ºC within the last 24 hours?**

🞎 No 🞎 Yes

**Did the patient have leucocytes ≤4,000 or ≥12,000 within the last 24 hours?**

🞎 No 🞎 Yes

**Did the patient present with purulent tracheal secretions?**

🞎 No 🞎 Yes

**Was there a positive culture?**

🞎 No 🞎 Yes

**If the culture test yields a positive resul:**

**How many pathogens are present?**____________________________

**What is the name of the pathogen?**____________________________

**Has the patient received oxygen therapy in the last 24 hours?** (e.g., nasal cannula, mask, venturi mask, non-invasive ventilation, invasive ventilation)

🞎 No 🞎 Yes

**For patients using oxygen therapy (cannula, mask) or ventilation, has oxygen saturation changed in the last 24 hours?**

🞎 No

🞎 Yes, saturation < 92%

🞎 Yes, saturation > 96%

**Has the patient received vasoactive drugs (e.g., adrenaline, dobutamine, dopamine, noradrenaline, vasopressin) in the last 24 hours?**

🞎 No 🞎 Yes

**Has the patient undergone dialysis in the last 24 hours?**

🞎 No 🞎 Yes

**Has the patient received continuous sedation with benzodiazepines (e.g., diazepam, midazolam) in the last 24 hours?**

🞎 No 🞎 Yes

**Was there a reduction or interruption in sedation in the last 24 hours?**

🞎 No 🞎 Yes

**Is prophylaxis for venous thromboembolism indicated?**

🞎 No 🞎 Yes

**Is the patient restricted to bed most of the time?**

🞎 No 🞎 Yes

**Does the patient have active bleeding?**

🞎 No 🞎 Yes

**Does the patient have an active peptic ulcer?**

🞎 No 🞎 Yes

**Does the patient have uncontrolled arterial hypertension (> 180/110 mmHg)?**

🞎 No 🞎 Yes

**Does the patient have coagulopathy (e.g., thrombocytopenia or INR > 1.5)?**

🞎 No 🞎 Yes

**Does the patient have an allergy or thrombocytopenia caused by heparin?**

🞎 No 🞎 Yes

**Does the patient have renal failure (creatinine clearance < 30 mL/min)?**

🞎 No 🞎 Yes

**Has the patient undergone cranial or ocular surgery within the last two weeks?**

🞎 No 🞎 Yes

**Has the patient undergone a cerebrospinal fluid collection in the last 24 hours?**

🞎 No 🞎 Yes

**Is the patient receiving prophylaxis for venous thromboembolism (e.g., heparin, enoxaparin)?**

🞎 No

🞎 Yes

🞎 Yes, on oral anticoagulant therapy

**What is the primary type of diet the patient is receiving?**

🞎 Total fasting

🞎 Oral diet

🞎 Enteral feeding

🞎 Parenteral nutrition

**Was at least one blood glucose measurement performed in the last 24 hours?**

🞎 No 🞎 Yes

**Did the patient experience hypoglycemia or hyperglycemia in the last 24 hours?**

🞎 No

🞎 Yes, blood glucose < 70 mg/dL (hypoglycemia)

🞎 Yes, blood glucose > 180 mg/dL (hyperglycemia)

**Does the patient currently have a central venous catheter?**

🞎 No 🞎 Yes

**Is there, or has there been, suspicion of a central venous catheter-related infection?**

🞎 No 🞎 Yes

**Has the patient been diagnosed with a central line-associated bloodstream infection (CLABSI)?**

🞎 No 🞎 Yes

**If the patient has CLABSI, was a new central line inserted?**

🞎 No 🞎 Yes

**If the patient has CLABSI, on the day or the day before the positive blood cultures were collected, did the patient have a central venous catheter (CVC)? (Includes central venous catheters, pacemaker introducers, pulmonary artery catheters, and peripherally inserted central catheters)**

🞎 No 🞎 Yes

**If the patient has CLABSI, is there a recognized pathogen growing in one or more blood cultures that is not related to an infection at another site?**

🞎 No 🞎 Yes

**If the patient has CLABSI, has a common skin contaminant grown in the blood culture?**

🞎 No 🞎 Yes

**If the patient has CLABSI, does the patient have at least one of the following symptoms: fever (>38°C), chills, or hypotension?**

🞎 No 🞎 Yes

**If the patient has CLABSI, are the symptoms unrelated to an infection at another site?**

🞎 No 🞎 Yes

**If the patient has CLABSI, has the same common contaminant grown in at least two blood cultures collected at separate times?**

🞎 No 🞎 Yes

**Does the patient use an indwelling urinary catheter?**

🞎 No 🞎 Yes

**Is there or has there been suspicion of a catheter-associated urinary tract infection (CAUTI)?**

🞎 No 🞎 Yes

**If the patient has CAUTI, was the catheter removed?**

🞎 No 🞎 Yes

**If the patient has CAUTI, did the patient have an indwelling urinary catheter on the day or the day before the positive urine cultures were collected?**

🞎 No 🞎 Yes

**If the patient has CAUTI, does the patient exhibit at least one of the following symptoms without another identified cause: fever (>38°C), increased frequency, urgency, dysuria, or suprapubic pain?**

🞎 No 🞎 Yes

**If the patient has CAUTI, does the urine culture show positive results with ≥100,000 microorganisms/mL (or 100,000 CFU/mL) involving no more than two species?**

🞎 No 🞎 Yes

**Is there a documented limitation of advanced life support in the patient’s medical record?** **(Includes dialysis, intubation, vasoactive drugs, or cardiopulmonary resuscitation)**

🞎 No 🞎 Yes

**Was the patient mobilized out of bed (e.g., walked or sat in a chair) in the last 24 hours?**

🞎 No 🞎 Yes

**In the last 24 hours, was the patient exposed to any form of mobilization?** **(e.g., passive limb movement in bed, sitting at the bedside or in a chair, walking)**

🞎 No 🞎 Yes

**In the last 24 hours, did the patient sit outside the bed (in a chair) or at the bedside with legs dangling and unsupported?**

🞎 No 🞎 Yes

**Is the patient currently on antibiotics?**

🞎 No 🞎 Yes

**If the patient is currently on antibiotics, provide the following details for each antibiotic:**

**Start date:**____________________________________________________

**End date:**____________________________________________________

**Name of the antibiotic:**_________________________________________

**ICU DISCHARGE FORM**

**ICU discharge date and time**:_______________________________________

**Vital status at ICU discharge**

🞎 Alive

🞎 Death

**Destination at ICU discharge**

🞎 Discharge to home

🞎 General ward

🞎 Step-down unit

🞎 Another ICU within the same hospital

🞎 Hospital transfer

**ICU READMISSION FORM**

**Was the patient previously admitted to this ICU during the current hospital stay and already included in the TELESCOPE2 study?**

🞎 No 🞎 Yes

**How many readmissions to the ICU occurred during this hospital stay?**___

**Did this readmission occur within 48 hours of ICU discharge?**___________

**What is the date of the ICU readmission?**____________________________

**HOSPITAL DISCHARGE FORM**

**What is the date and time of the hospital discharge?**___________________

**What was the patient's vital status at the time of hospital discharge?**

🞎 Alive

🞎 Death

**Was this hospital discharge a transfer to another hospital?**

🞎 No 🞎 Yes
